# Mapping brain lesions to conduction delays: the next step for personalized brain models in Multiple Sclerosis

**DOI:** 10.1101/2024.07.10.24310150

**Authors:** C. Mazzara, A. Ziaeemehr, E. Lopez Troisi, L. Cipriano, M. Angiolelli, M. Sparaco, M. Quarantelli, C. Granata, G. Sorrentino, M. Hashemi, V. Jirsa, P. Sorrentino

**Affiliations:** Department of Promoting Health, Maternal-Infant. Excellence and Internal and Specialized Medicine (PROMISE) G. D’Alessandro, University of Palermo, Palermo, Italy; Institute of Biophysics, National Research Council, Palermo, Italy; Aix Marseille Univ, INSERM, INS, Inst Neurosci Syst, Marseille, France; Institute of Applied Sciences and Intelligent Systems, National Research Council, Pozzuoli, Italy; Department of Medical Motor and Wellness Sciences, University of Naples “Parthenope”, Naples, Italy; Unit of Nonlinear Physics and Mathematical Models, Department of Engineering, Campus Bio-Medico University of Rome, 00128, Italy; Department of Advanced Medical and Surgical Sciences, University of Campania Luigi Vanvitelli, Caserta, Italy; Biostructure and Bioimaging Institute, National Research Council, Naples, Italy; Department of Economic, Legal, Informatics and Motor Sciences, University of Naples Parthenope, Nola, Italy; ICS Maugeri Hermitage Napoli, Via Miano 69 – 80145; University of Sassari, Department of Biomedical Sciences, Viale San Pietro, 07100, Sassari, Italy

**Keywords:** Multiple Sclerosis, Personalized brain models, conduction delays, Magnetoencephalography, Bayesian inference

## Abstract

Multiple sclerosis (MS) is a clinically heterogeneous, multifactorial autoimmune disorder affecting the central nervous system. Structural damage to the myelin sheath, resulting in the consequent slowing of the conduction velocities, is a key pathophysiological mechanism. In fact, the conduction velocities are closely related to the degree of myelination, with thicker myelin sheaths associated to higher conduction velocities. However, how the intensity of the structural lesions of the myelin translates to slowing of nerve conduction delays is not known. In this work, we use large-scale brain models and Bayesian model inversion to estimate how myelin lesions translate to longer conduction delays across the damaged tracts. A cohort of 38 subjects (20 healthy and 18 with MS) underwent MEG and MRI acquisitions, with detailed white matter tractography analysis. We observed that MS patients consistently showed decreased power within the alpha frequency band (8-13Hz) as compared to the healthy group. We also derived a lesion matrix indicating the percentage of lesions for each tract in every patient. Using large-scale brain modeling, the neural activity of each region was represented as a Stuart-Landau oscillator operating in a regime showing damped oscillations, and the regions were coupled according to subject-specific connectomes. We propose a linear formulation to the relationship between the conduction delays and the amount of structural damage in each white matter tract. Dependent upon the parameter *γ*, this function translates lesions into edge-specific conduction delays (leading to shifts in the power spectra). Using deep neural density estimators, we found that the estimation of *γ* showed a strong correlation with the alpha peak in MEG recordings. The most probable inferred *γ* for each subject is inversely proportional to the observed peaks, while power peaks themselves do not correlate with total lesion volume. Furthermore, the estimated parameters were predictive (cross-sectionally) of individual clinical disability. This study represents the initial exploration showcasing the location-specific impact of myelin lesions on conduction delays, thereby enhancing the customization of models for individuals with multiple sclerosis.

## Introduction

Multiple sclerosis (MS) is a chronic autoimmune disease that affects the central nervous system, provoking lesions that result in a wide variety of physical and cognitive disabilities, including paresthesia, vision loss, ataxia, mental changes and weakness (Gaby, 2013; Ghasemi et al., 2017). The etiology of MS is still unclear; nonetheless, it is understood that it is multifactorial and consist of complex interactions among different genetic and environmental factors (Bernard and de Rosbo, 1992; Filippi et al., 2018; Ghasemi et al., 2017; Yamout and Alroughani, 2018).The pathogenesis of MS entails a cascade of events in the immune system which result in inflammation, nerve demyelination, axonal/neuronal damage, and death of neuronal cells (Ghasemi et al., 2017; Miller, 2012).

Attacks on the myelin play a pivotal role in MS pathophysiology. Damage to the myelin, such as demyelination or decompaction, can lead to impaired conduction with lower velocities (Fowler and Gilliatt, 1981; Gutiérrez et al., 1995; Reutskiy et al., 2003). In fact, the conduction velocity of action potentials along the white matter bundles is proportional to the degree of myelination (Gutiérrez et al., 1995). Therefore, the thickness of the myelin sheath, which is also closely associated to the axon diameter, plays a significant role in determining conduction velocity (Gillespie and Stein, 1983; Ritchie, 1982; Sanders and Whitteridge, 1946).

Along these lines, characterizing conduction velocity and delays might be an effective way to estimate subtle damage to the myelin that, while contributing to the clinical picture, might not yet manifest as overt structural changes and, hence, might not be detected by structural imaging alone. Evoked potentials have been used to measure conduction velocity on selected white matter tracts, but assessing the delay across the whole brain is not feasible (Leocani et al., 2000). However, the overall dynamics, which depends on all the delays, is clinically relevant(Sorrentino et al., 2022). Recent studies have taken a stride forward in evaluating conduction delays and velocity though structural magnetic resonance imaging (MRI) (Drakesmith and Jones, 2019; Mancini et al., 2021), providing a non-invasive alternative. Mancini (Mancini et al., 2021) and Drakesmith (Drakesmith and Jones, 2019) leveraged MRI-derived microstructural measures to estimate conduction velocity and delays distributions across the whole brain, using features such as the axonal diameter and the myelin content (calculated through the g-ratio). However, there are still limitations, such as the MRI resolution limit, which can affect the ability to accurately measure microstructural parameters, hindering for example the complete characterization of axonal diameter distribution and, consequently, leading to estimation errors, especially for smaller axons. Crucially, how exactly the myelination, as estimated by the g-ratio, maps onto the conduction velocities remains unknown. One might expect that the relation between the g-ratio and the actual conduction velocities might change as a function of the edge considered. However, the validation of MRI measures is limited (Mancini et al., 2021). Furthermore, accurate estimates of the conduction velocities are generally not obtainable in scenarios characterized by low axon density (Drakesmith and Jones, 2019).

Deriving the average conduction velocity across the entire brain poses challenges mainly due to the lack of efficient model inversion algorithms operating at such a large scale (Hashemi et al., 2021, 2020). To overcome this difficulty, Sorrentino et al.(Sorrentino et al., 2023)opted to infer patient-specific average conduction velocities by integrating diffusion tensor imaging and source-reconstructed magnetoencephalography (MEG) data into a personalized large-scale brain model. Such personalized models were then inverted using a simulation-based inference (SBI) scheme with a class of deep neural networks known as neural density estimators (Gonçalves et al., 2020; Hashemi et al., 2023; Liu et al., 2021) to estimate the most likely conduction velocities.

In this paper, we hypothesize that a specific relationship exists between the extent of the myelin lesions, as measured from structural imaging, and the consequent rise in the conduction delays across the brain edges (Fig.1). To test for the existence of such relationship, and to invert it at the patient-specific level, we used the neural density estimators in each participant at the whole-brain level.

As explained in more detail in Sorrentino et al.(Sorrentino et al., 2023) each subject underwent both MEG and MRI, including white matter tractography analysis. Additionally, we obtained a lesion matrix describing the percentage of lesion for each edge in each patient. We used a large-scale brain models where each region of interest (ROI) is represented as a Stuart-Landau oscillator in a regime with damped oscillations. For each subject, we utilized the shift in the power spectrum to quantify the effect of the structural damage, measured in each white-matter bundle, on the conduction delays across these edges.

**Fig. 1.**
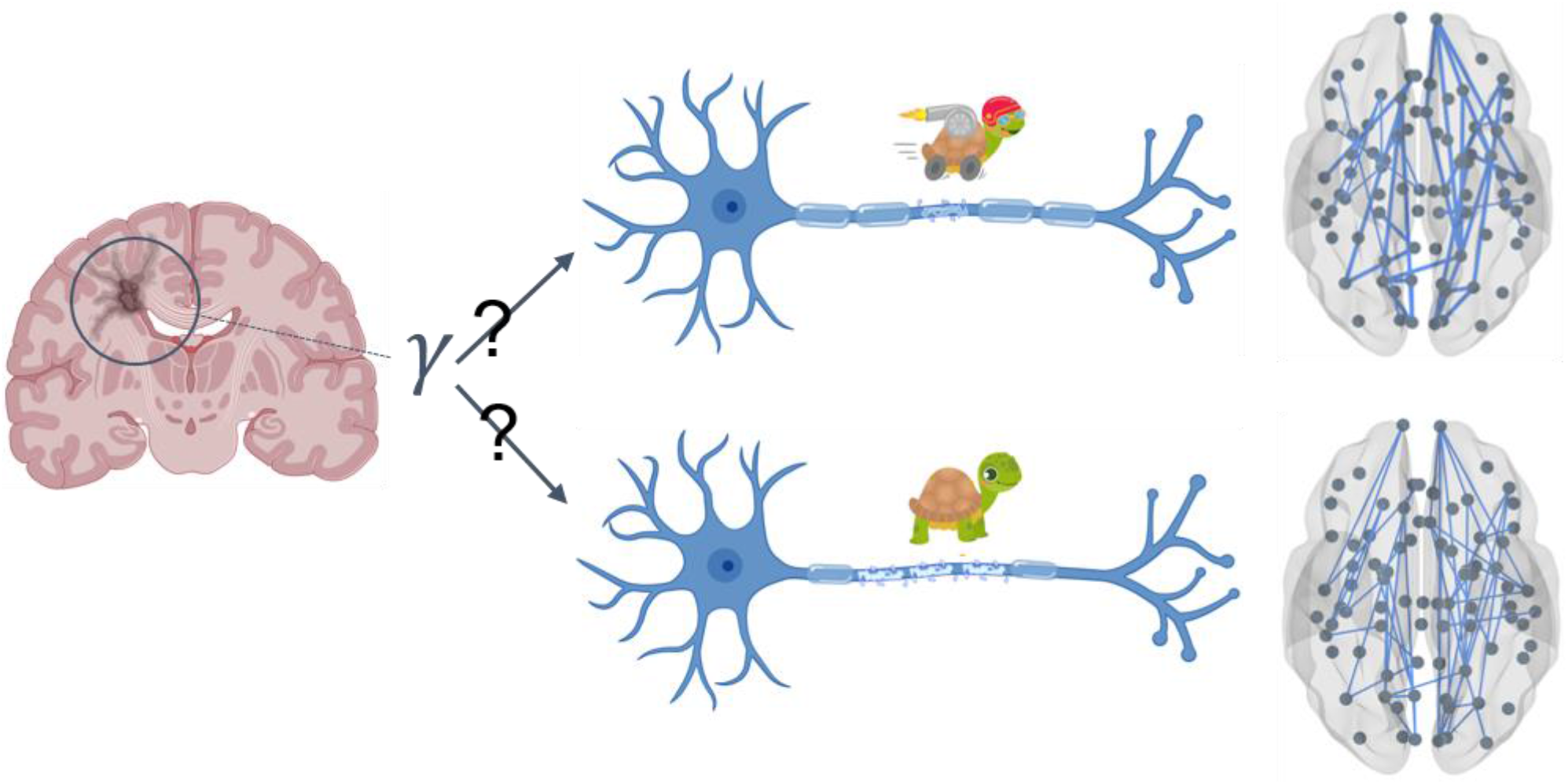
Model Overview: Gamma Parameter. On the left, the diagram illustrates a typical lesion in multiple sclerosis (MS). The variable gamma (*γ*) in our model represents the relationship between the structural lesion and the corresponding slowing of the velocity in the propagation of the nervous signal. The value of gamma gauges the severity of the effect of the lesion. Although the signal transmission is slower in patients as compared to the controls, the slowing can vary greatly. At the top of the diagram, less severe MS is depicted, characterized by fewer lesions that translate less prominently to higher delays, as represented by the thicker connections between the nodes. At the bottom, a more severe condition is represented, with more myelin lesions, severely slowed conduction velocities and, in the whole brain model, thinner connections.

## Results

We confirmed the presence of a shift in the alpha frequency peak and a reduction in alpha band (8-13 Hz) power spectra in 18 MS patients compared to 20 controls (Fig.2a). Then, we aimed to establish a relationship between the degree of lesion in specific edges (which is expected to affect conduction velocities) and the power spectrum of MEG recordings. To this end, subject-specific tractography was utilized to generate a large amount of synthetic data while varying the global coupling (scaling the structural connectivity matrix) and the parameter *γ*, that indicates the degree of influence of the lesions on the delay, from physiologically plausible ranges.Subsequently, low dimensional spectral features, including the area under the power spectral density (PSD), the amplitude of the peak frequency, and the peak frequency itself, were extracted from the synthetic dataset. Then we train neural density estimators to learn an invertible map between the effect of the lesions on conduction delays (represented by the parameter *γ* in our model) and the resulting data features extracted from random simulations. Note that, in each patient, the parameter *γ* weighs a different set of edges, that is, those who are lesioned in that patient. Fig. 2b illustrates the power spectrum as a function of the frequency for various values of the parameter *γ* . This figure demonstrates that increasing the *γ* parameter, and thus assigning higher weights to lesions (resulting in greater impact on conduction delays) leads to more pronounced reduction in the amplitude of the alpha peak in the spectrum, and to a shift in its frequency. In Fig.2c, one can observe the changes in the amplitude and frequency of the peak, as well as the variations in the area under the PSD, calculated from the alpha spectrum as a function of *γ* . All these spectral features decrease as *γ* increases.

**Fig. 2.**
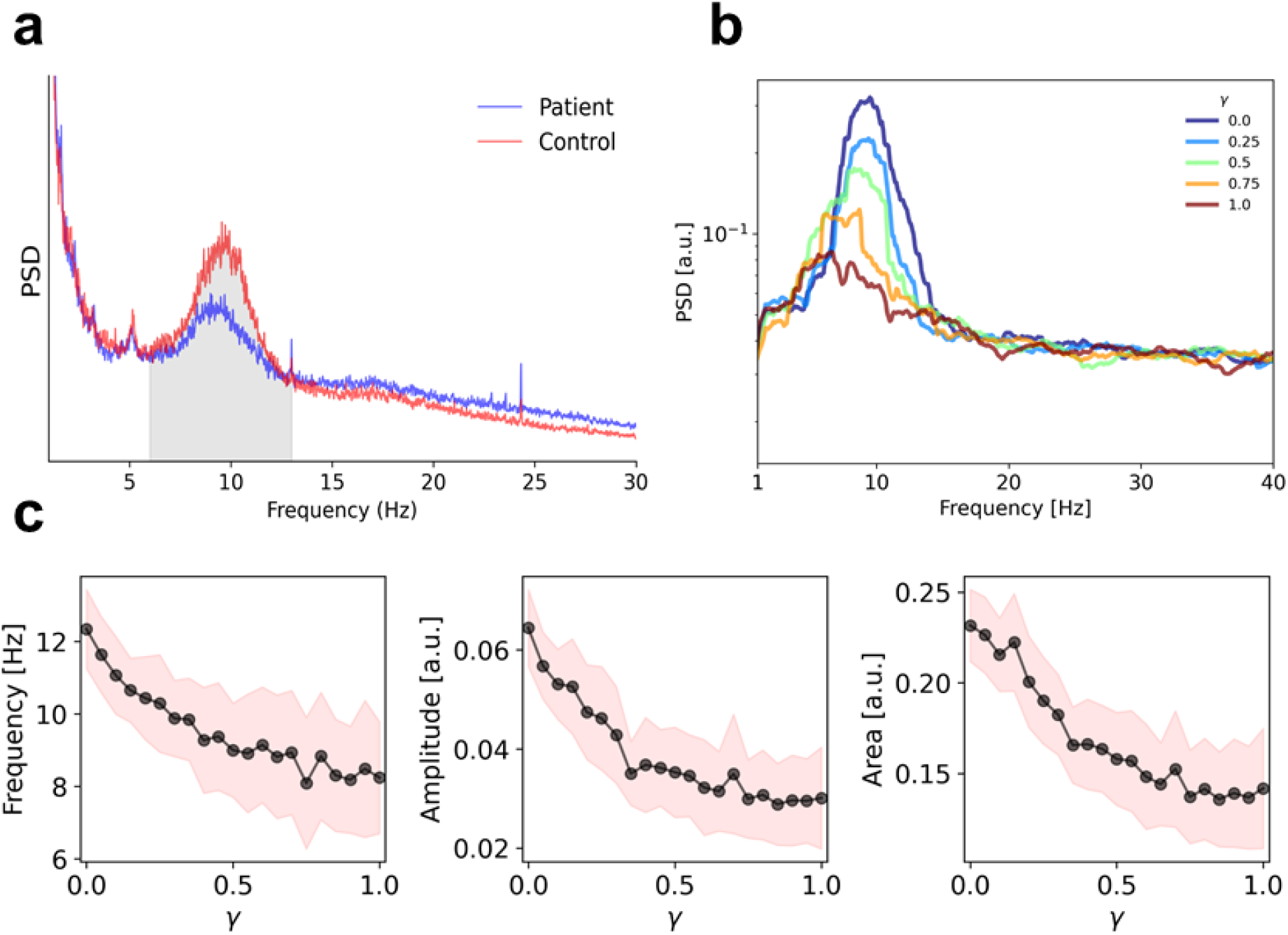
**a)** Median power spectral density for both controls (in red) and MS patients (in blue). **b)** Simulated Power Spectral Density (PSD) exhibits a peak in the alpha band, which decreases as the *γ* parameter increases. **c)** Peak frequency, Amplitude of peak frequency, and area under the PSD as a function of *γ* .

Then, we used SBI to build an invertible map between the values of *γ* (and G, i.e. the scaling factor for the connectome) to the corresponding data features. The validation of the SBI pipeline is shown in Fig.3. The neural density estimators (here, Masked Autoregressive Flows(Papamakarios et al., 2017)) are trained using 5000 random simulations. In Fig.3a shows the posterior distribution of *γ* given PSD features, while the parameters are drawn from a uniform prior distribution over the range [0,1]. For different model configurations, the SBI accurately estimates the posterior of parameter *γ* using PSD features. In Fig.3b, we showed different scenarios for *γ*, and we observed the accuracy of the estimated parameters versus the true values (with slope of 0.8 for the fitted maximum posteriors versus the true value).

**Fig. 3.**
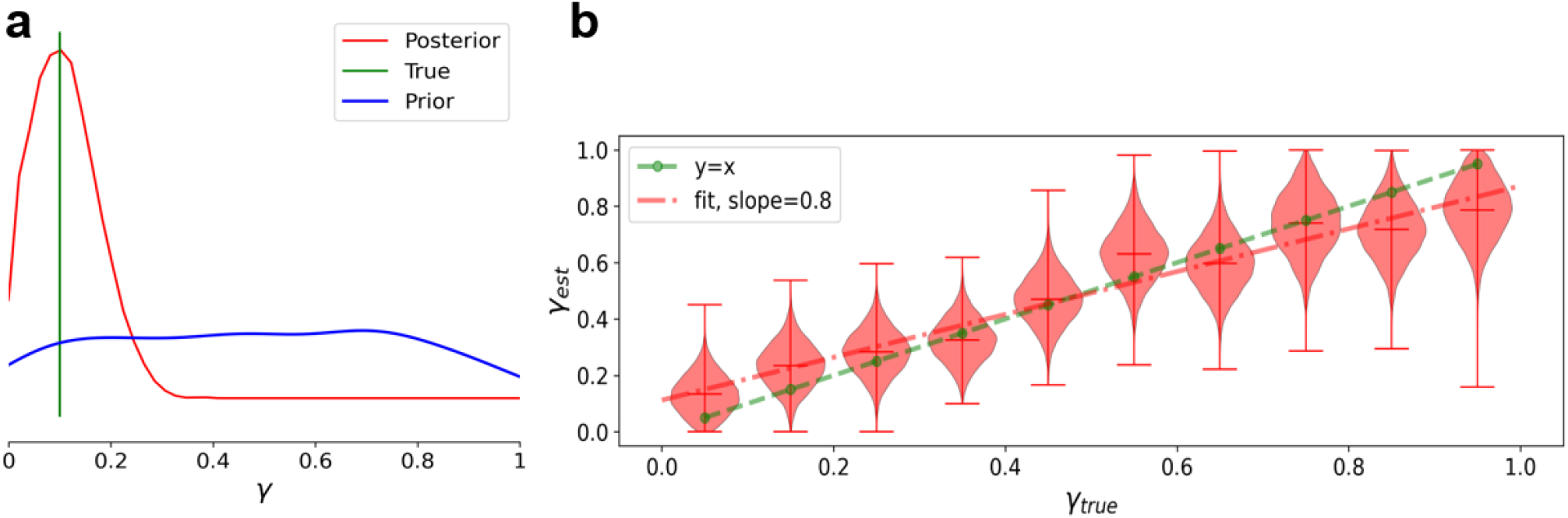
Validation of SBI using different model configurations over the parameter *γ*. **a)** Posterior distribution in red, True parameter in green, and the uniform prior distribution in blue for a synthetic diagnostic of the model inversion pipeline. SBI provides accurate recovery of the parameter *γ* . **b)** The violin plots in red show the estimated posterior densities by varying the parameter *γ*, the dashed red line represents a linear regression on the maximum values of the estimated posteriors; dashed green line represents the perfect fit (ground truth values). These results show the accuracy and consistency of the model inversion for different model configurations.

We invert this map and use it to estimate the most likely *γ*, in each individual subject, given the empirically observed PSD features. We show that the most plausible *γ* values for the subjects relate, as expected, to the intensity of the peak in the alpha frequency range (Fig.4a) (r= -0.83, p=0.00). However, the lesion load itself does not relate to the amplitude of the alpha-peak nor to the *γ* parameter (r= -0.38, p= 0.12, r= 0.00, p= 0.9, respectively), showing that the weighting of the lesions, that is, the effect that they exert on the slowing of conduction velocities, is likely topography-dependent and, as such, patient specific.

**Fig. 4.**
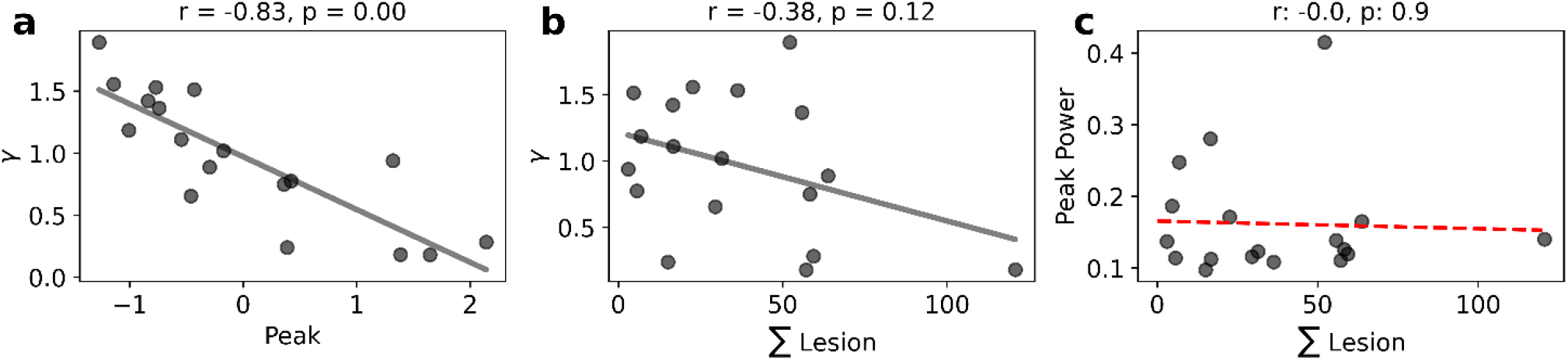
**a)** Correlation between *γ* and the amplitude of alpha-peak in MEG recordings (r= -0.83, p= 0.00). **b)** Correlation between *γ* and the lesion load (r= -0.38, p= 0.12). **c)** Correlation between the empirical peak and the lesion load (r= 0.00, p= 0.9).

Then we assessed the validity of the inferred individual *γ* values in predicting clinical disability. Using a multilinear model, we predicted clinical disability measured by the Expanded Disability Status Scale (EDSS), considering variables such as age, disease duration, total lesion volume, and inferred *γ* values as predictors. Adding *γ* to the predictive model enhances the predictive power on clinical disability, although without reaching statistical significance, as shown in Fig. S1. The model has been validated using a leave-one-out cross-validation scheme. Moreover, it is evident that the total lesion load has less predictive power. Although failing to reach statistical significance, the increase in the explained variance implies clinical relevance despite being non-predictive.

## Discussion

In MS, the patients consistently exhibit a decrease in power within the alpha frequency band (8-13 Hz), as reported by several studies. Cover et al.(Cover et al., 2006)discovered a notable reduction in interhemispheric synchronization in the alpha band, measured via MEG among MS patients, while Bsar-Eroglu (Bsar-Eroglu et al., 1993) observed significant reductions in both alpha band synchronization and amplitude. These findings were further supported by Schependom (Van Schependom et al., 2021),who linked the decrease in alpha power to increased brain atrophy and lesion load. Additionally, Sorrentino et al. (Sorrentino et al., 2023)identified substantial differences in alpha band features between MS patients and age-matched controls. Specifically, a shift in the power spectra of brain activity has been observed in MS, characterized by reduced power in the alpha frequency band and a shift in the peak frequency towards lower frequencies. This reduction in alpha power is believed to result from attacks by the immune system on the myelin, leading to a slowing down of nerve signal conduction, as evidenced by recent studies showing widespread delays throughout the brain in MS patients compared to healthy individuals (Berman et al., 2020; Sorrentino et al., 2022).

The main goal of this work is to estimate a relationship between the lesions specific to each patient and the resulting conduction delays, via the resulting changes in the power spectrum, particularly in the alpha band. We found a negative correlation between *γ*, a parameter that weighs the influence of the lesion mask, and the amplitude of the alpha peak in the spectrum. However, we also observe that the power peaks themselves do not correlate with the individual total lesion volume. All in all, this finding highlights the importance of the specific topography of lesions, since the impact of similar total lesion volumes onto the overall spectral features varies greatly across patients. An interesting hypothesis would be that each tract holds a specific way to impact the overall large-scale dynamics. In this case, lesions involving specific structures, such as the cortico-thalamic loop or the posterior areas, might more prominently affect the global dynamics. Along these lines, it was shown that thalamic atrophy in MS is linked to widespread disruption of cortical functional networks, which is associated with poorer cognitive and clinical outcomes in MS patients(“Functional brain networks,” n.d.). To test this hypothesis, we plan to investigate whether specific types of edges structural lesion translate affect conduction velocities in an edge-specific fashion, or if each edge presents a unique contribution. To this end, larger multimodal datasets are needed. This hypothesis, besides the neuroscientific interest, also holds practical relevance. In fact, inferring edge-specific delays is not feasible due to a parameter explosion. However, as we have seen, the specific topography of the lesions, that is, the specific edges that are damaged, plays a role to the final effect on the large-scale dynamics. In this context, our work provides a way to translate patient-specific lesion topographies to changes in the conduction velocities in all the edges involved, according to one rule. This way, we hope to lead to better personalization of large-scale models, while preventing this problem to become too high-dimensional.

In conclusion, developing and refining increasingly personalized models to monitor the therapeutic treatment of MS patients is crucial. The variability among individuals significantly influences treatment outcomes and tailor healthcare options to each patient’s specific needs is expected to enhance treatment outcomes (Jirsa et al., 2023, 2017; Lang et al., 2023; Wang et al., 2023). This is the first study demonstrating the topography-specific effect of myelin lesions on conduction delays, adding one step further to the personalization of models in individuals with MS.

## Methods

### Participants

Participants (both controls and patients) were recruited from the outpatient clinic of the Institute for Diagnosis and Cure Hermitage Capodimonte in Naples, Italy. Selection of patients with multiple sclerosis (MS) followed the 2017 McDonald criteria (Thompson et al., 2018), which excluded individuals under the age of 18, those with recent relapses or steroid treatments within three months, and those with a history of illicit drug, stimulant, amphetamine, barbiturate, and cannabinoid use or any other history of central nervous system disorders besides MS. Each patient underwent a comprehensive neurological examination, along with various assessments including the Expanded Disability Status Scale (EDSS). Table 1 provides a summary of the demographic data, key clinical characteristic, and primary radiologic findings for the MS cohort. Controls were chosen from caregivers and patients’ partners. Ethical approval was obtained from the local Ethics Committee (Prot.n.93C.E./Reg. n.14-17OSS).

**Table 1.** MS cohort features.

|  | Controls | MS | p-value |
| --- | --- | --- | --- |
| Age (years) | 45.8 (±11) | 44.9 (±) | 0.8 |
| Education (years) | 13.6 (±) | 13.8 (±) | 0.9 |
| Gender (m/f) | 6/14 | 6/12 | 0.3 |
| Disease duration (months) | - | 187.7 (±131.8) | - |
| EDSS | - | 4.5 (±1.9) | - |
| SDMT | - | 40.3 (±13) | - |
| FSS | - | 36.1 (±14) | - |
| BDI | - | 12.8 (±1.3) | - |
| LL | - | 12.959 (±12.253) mm<sup>3</sup> | - |

DSS,Expanded Disability Status Scale; SDMT, Symbol Digit Modalities; FSS, Fatigue Severity Scale; BDI BeckDepression Inventory; LL,lesion load.

### MRI acquisition and processing

The MRI data acquisition and processing followed the methods described in Sorrentino et al (Sorrentino et al., 2022). An MRI scanner operating at 1.5 Tesla (Signa, GE Healthcare) was utilized, with echo-planar imaging employed for DTI (Diffusion Tensor Imaging) reconstruction and 3D-FLAIR volume utilized for WM (White Matter) lesion segmentation. Preprocessing of the diffusion MRI data included correction for head movements and eddy current distortions, using software modules provided in the FMRIB software library (FSL; http://fsl.fmrib.ox.ac.uk/fsl). This step is crucial to ensure proper alignment and artifact-free data, mitigating disturbances caused by head movement or other sources during scanning. Additionally, a brain mask was generated from the B0 images using the Brain Extraction Tool routine.

Subsequently, a diffusion tensor model was fitted to each voxel, allowing quantification of water diffusion direction and anisotropy in biological tissues. Using the Fiber Assignment by Continuous Tracking (FACT) algorithm, fiber tracts representing major connectivity pathways in the brain were generated. Two cortical study-specific ROI datasets were obtained by masking ROIs available in the AAL and Desikan–Killiany– Tourville (DKT) atlases. Spatial normalization of each participant’s FA volume was performed to obtain patient-specific ROI sets.

Furthermore, MS lesion maps were generated by segmenting the 3D-FLAIR volume using the lesion prediction algorithm implemented in the Lesion Segmentation Tool (LST toolbox version 3.0.0;www.statistical-modeling.de/lst.html) for the Statistical Parametric Mapping software package (SPM). After obtaining the lesion map, the 3D-FLAIR volume was aligned (or “coregistered”) with another type of MRI sequence called EPI (Echo Planar Imaging) for the same patient. This coregistration process, described by Ashburner and Friston in 1997 (Ashburner and Friston, 1997), ensures that the images from different MRI sequences are spatially aligned with each other. Once the coregistration was completed, the transformation matrix used to align the 3D-FLAIR volume with the EPI sequence was applied to the MS lesion volume. This step effectively transferred the spatial information of the lesions onto the same coordinate space as the EPI sequence. To ensure accurate alignment, the WM lesion volume was resampled using a method called nearest-neighbor interpolation. This interpolation method involves assigning each new voxel in the resampled volume the value of the nearest voxel in the original volume, helping to preserve the integrity of the lesion information during the transformation process.

The resulting output of this process is a set of lesion masks specific to each patient, accurately aligned with the DTI volume. In this way, these lesion masks can be used for further analysis and investigation of the relationship between MS lesions and brain connectivity patterns obtained from DTI data.

Finally, the average length of fibers connecting each pair of ROIs and whether the voxels traversed by these fibers contained MS lesions were computed separately for the AAL and DKT ROI sets, utilizing an in-house routine written in Interactive Data Language (IDL). The length of each tract was calculated for each subject by averaging the physical distances covered by the fibers within the tract. These distances were obtained by summing the straight-line distances between the points where the fiber changes direction.

### MEG pre-processing and source reconstruction

Each participant underwent MEG recordings consisting of two eyes-closed resting-state segments, each lasting 3 minutes and 30 seconds. Electrooculogram (EOG) and electrocardiogram (ECG) signals were recorded to identify physiological artifacts such as eye blinks and heart activity. An anti-aliasing filter was applied to the MEG signals, acquired at 1024 Hz, assuring that the signal is properly sampled without distortion or loss of information. A fourth-order Butterworth IIR band-pass filter (0.5-48 Hz) was subsequently applied. MEG preprocessing and source reconstruction procedures followed the detailed protocols outlined in (Sorrentino et al., 2021). Preprocessing and source reconstruction operations made use of the Fieldtrip Toolbox. Principal component analysis and independent component analysis were employed to eliminate environmental noise and physiological artifacts. Source reconstruction followed the protocols described in (Sorrentino et al., 2022).

In summary, the signal time series were reconstructed using 84 Regions of Interest (ROIs) based on the DKT atlas. Utilizing the volume conduction model proposed by Nolte (Nolte, 2003), the reconstruction process involved applying the linearly constrained minimum variance (LCMV(Van Veen and Buckley, 1988)) beamformer to reconstruct the signal sources based on the centroids of each ROI.

### Design of the synthetic model

Whole-brain models are computational representations designed to mimic the intricate workings of the entire human brain by integrating its various regions as a network node, placing a dynamical model at each brain region. These models strive to simulate the brain as a unified entity, considering both its anatomical structure, the nonlinear dynamics at each brain regions, and the interactions among different brain regions. Their main objective is to deepen our understanding of how the brain processes information, generates consciousness, and performs cognitive functions, and notably dysfunctioning at various brain diseases (Jirsa et al., 2023; Wang et al., 2023). Over time, whole-brain models have proven invaluable in elucidating neuropathologies such as Alzheimer’s disease, schizophrenia, epilepsy, traumatic brain injury, and others (Pathak et al., 2022; Wang et al., 2024). Their clinical utility extends to providing prognostic tools and predictive insights for various neurological disorders. However, given the diverse origins and mechanisms of brain pathologies, these models are customized to suit the specific etiology of each disease.

To effectively capture the dynamics of our system and construct a realistic brain network model with accurate connectivity and temporal delays, we opted to represent each ROI using a Stuart-Landau oscillator (Cabral et al., 2022). The Stuart-Landau model offers simplicity and versatility, making it suitable for studying a wide range of dynamic systems, particularly those with temporal delays. By incorporating a term for temporal delay into Stuart-Landau’s differential equation, this model provides a robust mathematical framework for analyzing the impact of delays on system dynamics. In contrast to other models, the Stuart-Landau model explicitly accounts for temporal delays, enabling a more comprehensive understanding of their influence on overall system behavior over time.

The Stuart-Landau system exhibits two potential solutions: damped or limit cycle solutions, contingent upon the bifurcation parameter *a*. For *a* < 0, we will have damped oscillations, while for *a* > 0, limit cycle solutions. In this study, we used a *=* − 5 to reflect the dampened characteristics of local oscillations. As previously done in Sorrentino et al. (Sorrentino et al., 2023), oscillators within the system are interconnected via white matter, with coupling strength determined by subject-specific DTI fiber counts (*C*_*jk*_). The activity of each ROI is expressed as:

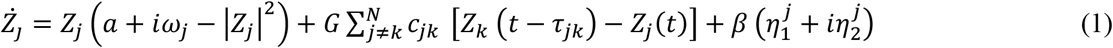

where:

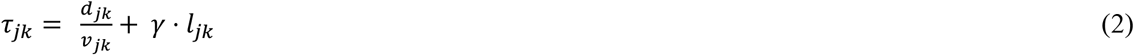

Each ROI functions as a local E-I (excitatory-inhibitory) unit oscillating in the gamma band, set at 40 Hz (*ω*_*jk*_). Parameter *G* is the global coupling parameter that represents the overall level of interconnection between the different parts of the system, influencing the strength of interactions among diverse brain regions.

*τ*_*jk*_ is the signal conduction time estimated by dividing inter-node Euclidean distances *d*_*jk*_ by the conduction velocity *v*_*jk*_ plus a contribution caused by the lesion. In this case, *τ*_*jk*_ depends on the degree of the lesion in every specific tract of the individual patient. In other words, the delay (specific for every tract) depends on *l*_*jk*_ that describes how damaged the tract in question is, while *γ* is a parameter representing the intensity of the influence of the lesions on the delay. Random noise β (mean=0, std=1e-4) is introduced to each oscillator to mimic stochastic variations.

### Model inversion

Inference is the top-down method of drawing conclusions or making predictions based on observations. In other words, it is the process of advancing from initial premises and observations to logical conclusions by quantifying the uncertainty in a model, providing an objectification of the validity of the decision (Jirsa et al., 2023).

Bayesian approach provides the posterior probability of an event, given a set of observed data. Bayes theorem states that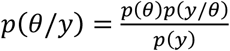, where *p*(*θ*/*y*) is the probability of a hypothesis given the data (known as the posterior probability), *p*(*θ*) is the probability of the parameter before seeing any data (the probability of the hypothesis itself, known as the prior probability), *p*(*y*/*θ*) is the probability of data given the hypothesis (known as the likelihood) and p(y) is the probability of the data independently from any parameter (known as model evidence). This means that we can update our prior knowledge, represented by prior probabilities with new information provided by observed data to refine our estimates about parameters of interest, incorporating in this way both background knowledge and new empirical evidence to obtain a more accurate estimation of quantities of interest. In this study, the unknown parameter set is *θ =* {*γ*, *G*}, where *γ* is the weight of the lesion load in a specific patient and *G* is the coupling scaling factor, whereas *y* represents the PSD features extracted from the MEG data of each patient.

Efficient Bayesian inference faces challenges in evaluating the likelihood function (*p*(*y*/*θ*)) given samples from the prior distribution. Particularly for whole-brain scales, computational costs make likelihood-based approaches sampling impractical. Simulation-based Inference (SBI; (Cranmer et al., 2020)), or likelihood-free inference, offers a solution by using low-dimensional data features, efficiently performing Bayesian inference for high-dimensional and complex models where calculating the likelihood function is intractable. In this case, the simulator can be used as a black-box whose internal workings do not accessible and are not required to be differentiable, but it can generate synthetic data similar to the observed data (Hashemi et al., 2023). To efficiently conduct SBI, a class of deep artificial neural networks called Normalizing-Flows (Rezende and Mohamed, 2015) can be used to learn an invertible transformation between parameters and data features from a set of simulations with parameters drawn from prior distribution.

We first validate SBI on the whole-brain model of Stuart-Landau oscillators using synthetic data generated with subject-specific structural connectomes by recovering ground-truth parameters, then apply it to empirical MEG data. For the training (here using Masked Autoregressive Flows; (Papamakarios et al., 2017)), the parameters are drawn from a uniform prior in the range *γ* ∈ U(0,1), with the budget of 5000 simulations. The set of low dimensional data features consists of the frequency, the peak amplitude of the mediated power spectrum, and the area under the curve in alpha band.

## Data Availability

The MEG data and the reconstructed avalanches are available upon request to the corresponding author (Pierpaolo Sorrentino), conditional on appropriate ethics approval at the local site. In case data are requested, the corresponding author will request an amendment to the local ethical committee. Conditional to approval, the data will be made available.

## Acknowledgements

The work was supported by the Sanofi iDEA-TECH-Awards, the European Union’s Horizon 2020 research and innovation program under grant agreement No. 101147319 (EBRAINS 2.0 Project) and No. 101137289 (Virtual Brain Twin Project). We also acknowledge a contribution from the Italian National Recovery and Resilience Plan (NRRP), M4C2, funded by the European Union - NextGenerationEU (Project IR0000011, CUP B51E22000150006, “EBRAINS-Italy”).

