## Supplemental Fig.S1 for "Mapping brain lesions to conduction delays: the next step for personalized brain models in Multiple Sclerosis"

### Supplementary Materials

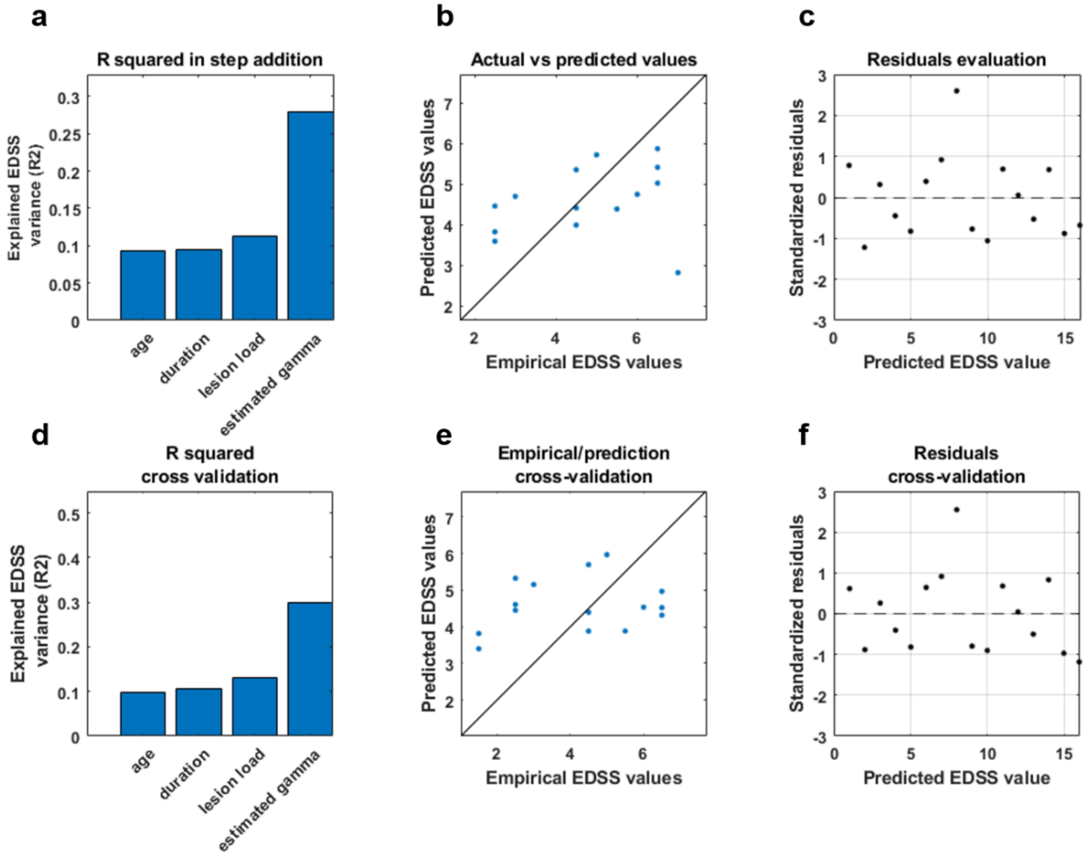


**Fig.S1 Clinical outcome prediction. a-d** Variance explained by the model adding 4 predictors: age, duration, lesion load and estimated γ. The parameter γ enhances prediction accuracy in both classical multilinear ($R2=0.2793$; $AdjR2=0.017226$) (age $\beta=0.0227$ $\rho=0.7328$; duration $\beta=0.0015$ $\rho=0.7547$; lesion load $\beta=-0.0000$ $\rho=0.5921$; γ $\beta=-3.3099$ $\rho=0.1366$) and cross-validated models ($R2=0.29882$; $AdjR2=0.018348$) (age $\beta=0.0327$ $\rho=0.7229$; duration $\beta=0.0011$ $\rho=0.7421$; lesion load $\beta=0.0000$ $\rho=0.5968$; γ $\beta=-3.3972$ $\rho=0.1784$). **b-e** Predicted versus empirical EDSS scores; **c-f** Residuals evaluation.
